# Age Estimation from Blood Test Results Using a Random Forest Model

**DOI:** 10.1101/2024.02.06.24302114

**Authors:** Satomi Kodera, Osamu Yokoi, Masaki Kaneko, Yuka Sato, Susumu Ito, Katsuhiko Hata

## Abstract

**Background and Objectives:** From the perspective of preventive medicine, in situations where screening tests are widely used, this study aims to clarify the role of screening data on ageing and health problems by estimating age from screening data with verifying the number of data items required.

**Materials and Methods:** A Python random forest model was generated using Chat GPT and tested.

**Results:** When using all 71 items, including gender, for the test results, a high accuracy of R^2^ = 0.7010 was obtained when there were 9243 training data sets (80% of the total number of data sets). The R2 decreased slightly to 0.6937 when the number of data items was reduced to 15 by discarding lesser importance items. When the number of data sets were less than 800 or when the number of data items were less than 7, the R^2^ value fell below 0.6. Interestingly, a higher age was tended to be estimated for post-menopausal women compared to pre-menopausal women.

**Conclusions:** The age estimated from blood data by the random forest model (blood age, so to speak) is so precise that it can be useful for assessing physical ageing state. However, the specific relationship between blood age and health status is still unclear, waiting for future research in order to deepen our understanding of this area.

## 1. Introduction

The Random Forest model is a machine-learning algorithm devised by Leo Breiman that uses multiple decision trees to make predictions by majority rule [1-4]. Random forests can be used for classification, regression and clustering.

In recent years, various blood items have been widely used as screening tests. Reference ranges and clinically defined values have been established for each of these test items, but efforts to combine multiple items for a comprehensive assessment, considering correlations, are still uncommon. This is partly due to the non-linear nature of the relationships between test items, making it difficult to create indices combining them.

Advances in machine learning, however, have facilitated non-linear multivariate analysis, allowing predictions based on the results of multiple test items. In particular, machine learning models, known as random forests, have attracted attention for their accuracy, resistance to overtraining, assessment of the importance of features, ease of parameter tuning and speed-up due to parallel processing [5-15].

In this study, we utilized ChatGPT to generate the Python code for random forest regression. With ChatGPT, highly accurate Python code can be generated in a short time, even without in-depth knowledge of machine learning [16, 17]. In addition, ChatGPT is based on natural language processing, which makes it intuitive and easy to communicate code generation requirements, enabling rapid prototyping and code modification.

This study introduces an approach to estimate age from test results, referred to as ‘blood age’, using Python code that implements a random forest model generated by ChatGPT. The aim is to investigate its potential as a new screening marker by taking into account its difference from actual age. Frailty Syndrome is an age-related deterioration of health status, so the test results should suggest the extent of health status deterioration, and the question of how age can be inferred from tests should be interesting from this perspective.

## 2. Materials and Methods

Target population: the study included 11554 men and women aged 0-95 years who underwent screening tests (60 blood tests, 8 urine tests and 2 saliva tests) from February 2020 to August 2023. Age and gender distribution are presented in Table 1.

**Table 1.** Age and gender distribution.

| Age (y.o.) | Number(%) |  |  | % of female |
| --- | --- | --- | --- | --- |
|  | Total | Male | Female |  |
| 0-9 | 45 ( 0.39 ) | 22 ( 0.72 ) | 23 ( 0.27 ) | 51.11 |
| 10-19 | 568 ( 4.92 ) | 297 ( 9.76 ) | 271 ( 3.18 ) | 47.71 |
| 20-29 | 947 ( 8.20 ) | 265 ( 8.71 ) | 682 ( 8.01 ) | 72.02 |
| 30-39 | 1839 ( 15.92 ) | 436 ( 14.33 ) | 1403 ( 16.48 ) | 76.29 |
| 40-49 | 2643 ( 22.88 ) | 574 ( 18.86 ) | 2069 ( 24.31 ) | 78.28 |
| 50-59 | 2306 ( 19.96 ) | 569 ( 18.70 ) | 1737 ( 20.41 ) | 75.33 |
| 60-69 | 1615 ( 13.98 ) | 446 ( 14.66 ) | 1169 ( 13.74 ) | 72.38 |
| 70-79 | 1207 ( 10.45 ) | 342 ( 11.24 ) | 865 ( 10.16 ) | 71.67 |
| 80-89 | 367 ( 3.18 ) | 90 ( 2.96 ) | 277 ( 3.25 ) | 75.48 |
| 90-99 | 17 ( 0.15 ) | 2 ( 0.07 ) | 15 ( 0.18 ) | 88.24 |
|  | 11554 | 3043 | 8511 |  |

The test items used in the study were as follows.

### Blood tests

total protein (TP), albumin/globulin ratio (A/G), albumin (ALB), alpha 1 glob-ulin (A1G), alpha 2 globulin (A2G), beta 1 globulin (B1G), beta 1 globulin %(B1G%), beta 2 globulin (B2G), beta 2 globulin (B1G%), gamma globulin (GG), total bilirubin (T-B), direct bilirubin (D-B), aspartate transaminase (AST), alanine transaminase (ALT), alkaline phosphatase (ALP), lactate dehydrogenase (LDH), cholinesterase (CHE), γ-GTP, amylase (AMY), creatine phosphokinase (CPK), blood sugar (BS), HBA1c, C-reactive protein (CRP), ferritin, total cholesterol (TC), free cholesterol, Cholesterol ester ratio, HDL cholesterol, LDL cholesterol, triglycerides (TG), arterial stiffness index (AI), free fatty acids (FFA), uric acid (UA), urea nitrogen (BUN), creatinine (CRE), sodium (Na), potassium (K), chlor (Cl), calcium (Ca), magnesium (Mg), copper (Cu), zinc (Zn), zinc/copper ratio (Zn/Cu), serum iron (Fe), unsaturated iron binding capacity (UIBC), white blood cell count (WBC), red blood cell count (RBC), haemoglobin (HB), haematocrit (HT), mean corpuscular volume (MCV), mean corpuscular HB volume (MCH), HB concentration (MCHC), platelet count (PLT), reticulocyte (RET), basophil (Baso), eosinophil (Eosino), neutrophil (Neut), lym-phocyte (Lymph), monocyte (Mono), 25-OH vitamin D (25-OH-VD).

### Urinalysis

urine sugar (uGlu), urine protein (uP), urine urobilinogen (uUB), urine occult blood reaction (uOBR), urine specific gravity (uSG), urine pH (uPH), urine bilirubin (uB), urine ketone bodies (uKB).

(-) to -1, (+-) to 0, (+) to 1, (2+) to 2, (3+) to 3

### Saliva test

salivary haemoglobin (sHb), salivary lactate dehydrogenase (sLD).

Missing values were less than 0.70 % of the total and were filled by the medians.

Urine sugar (uGlu), urine protein (uP), urine urobilinogen (uUB), urine occult blood reaction (uOBR), urine bilirubin (uB), urine ketone bodies (uKB) are Result values were converted as follows.

The dataset is divided into training and test data sets. This approach employs ‘supervised learning’, a method where models are trained on pre-labeled data and subsequently used to make predictions on new data [1]. The training data sets comprise 80% of the total dataset (9243 data sets), with the remaining 20% (2311 data sets) designated as test data sets. The analysis utilizes a Python-based random forest model. Data items include gender and 70 screening test items, with age set as the target variable. Based on this, the model was developed and trained. Subsequently, the estimated age from the test data sets was computed and compared to the actual age. The coefficient of determination, R^2^ (R squared), is used to evaluate model accuracy.

## 3. Results

The performance of the random forest model and the linear regression model in predicting age was compared (Fig. 1). The R^2^ value for age predicted using the random forest model was 0.7010 (Fig. 1A), whereas the R^2^ value for age predicted using the linear regression model was 0.5853 (Fig. 1B), indicating that the random forest model was more accurate. Based on these results, a detailed evaluation of the impact of the number of training data sets, the type of features required for reproducible age estimation, and the influence of gender differences and their associated characteristics on prediction performance was conducted in the random forest model.

**Figure 1.**
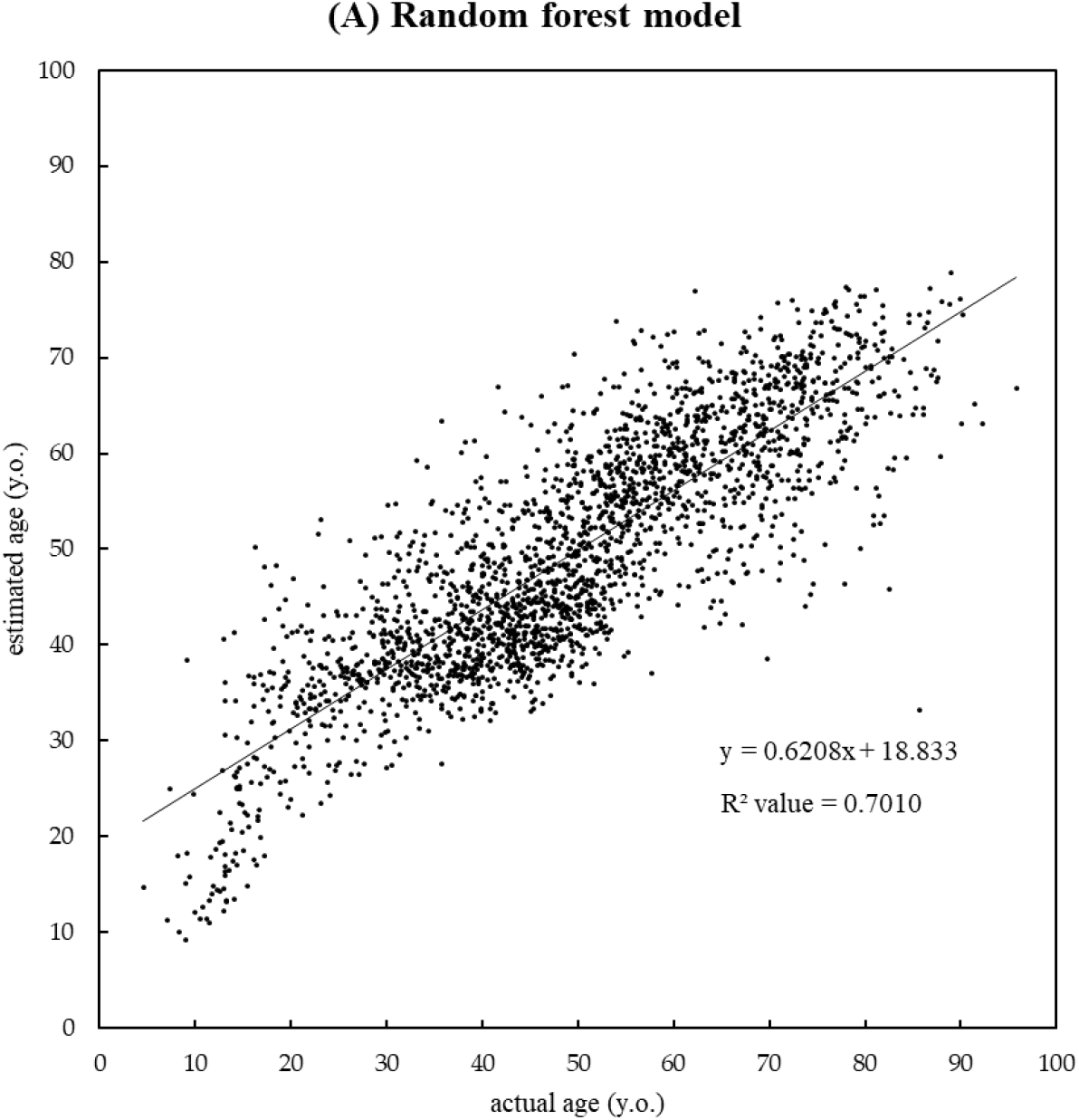
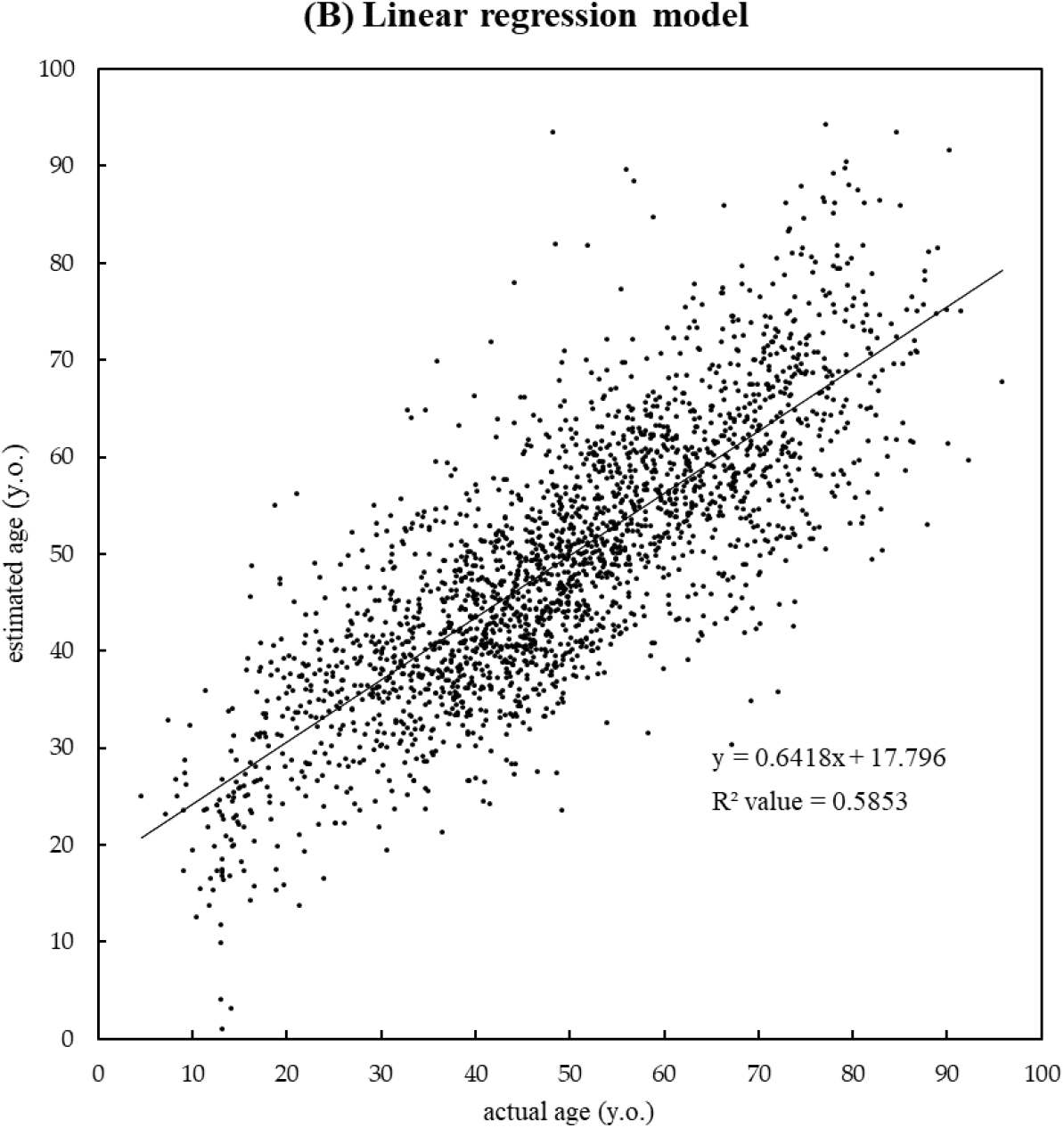
Comparison of the performance of the random forest model and the linear regression model in age prediction. (A) Relationship between the age estimated using the random forest model and the actual age (R^2^ value = 0.7010). (B) Relationship between the age estimated using the linear regression model and the actual age (R^2^ value = 0.5853).

### 3.1. Evaluation of age estimation by number of training data sets

The number of training data sets was incrementally increased from 100 to 10,000 in increments of 100 to assess the effect of the size of the training data set on the performance of the model (Fig. 2). As a result, it was observed that with 100 training data sets, the R^2^ value was less than 0.45, while with more than 800 training data sets, the R^2^ value improved to more than 0.60. Notably, the R^2^ values ranged from 0.68 to 0.69 for training data sets between 8,000 and 10,000, suggesting that training data sets with more than 8,000 are sufficient for accurate model estimation.

**Figure 2.**
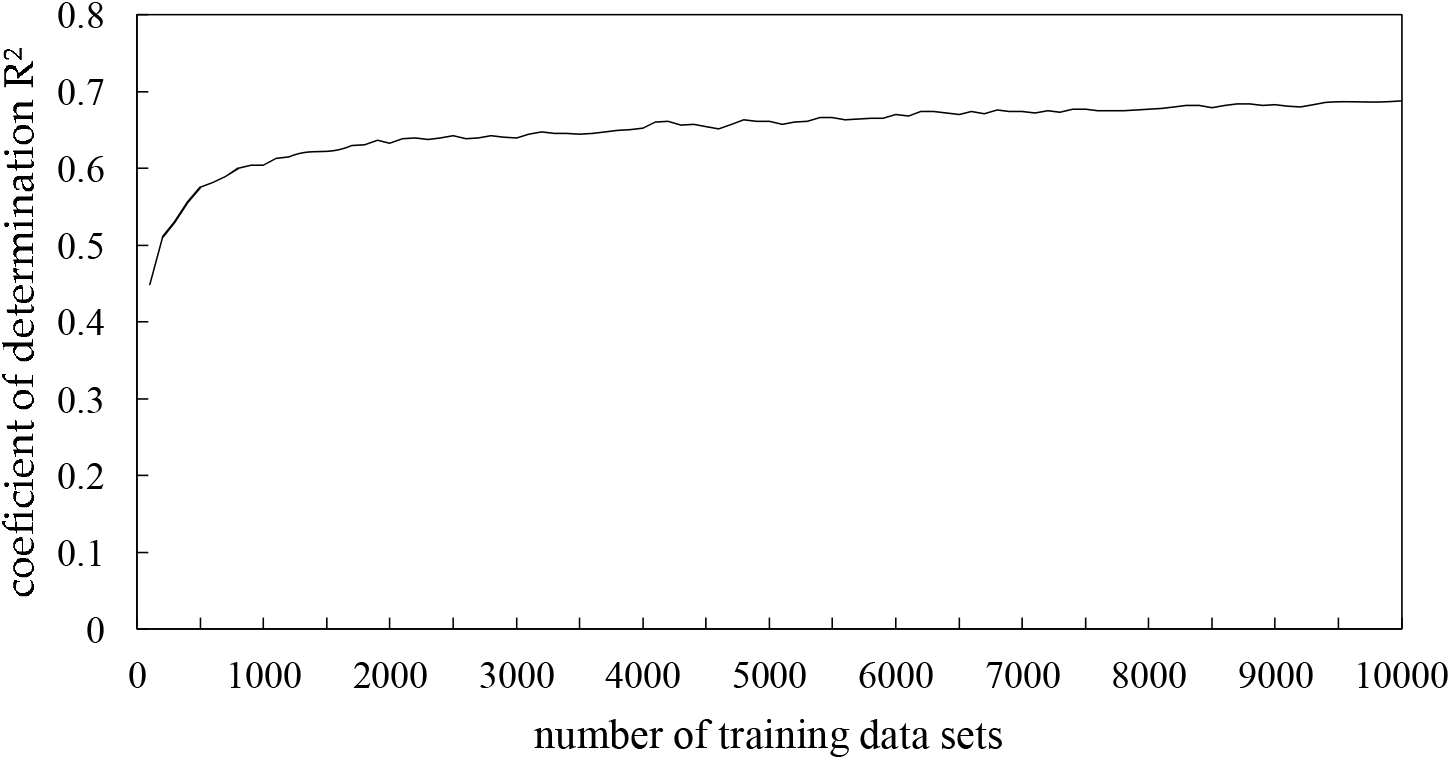
Relationship between the numbers of training data sets and coefficients of determination. The R^2^ value progressively increases with more training data sets. Initially, with 100 data sets, the R^2^ is below 0.45. It exceeds 0.60 with more than 800 data sets and stabilizes between 0.68 and 0.69 for 8,000 to 10,000 data sets.

### 3.2. Evaluation of age estimation with features

To investigate the usefulness of training using only blood tests, urine and saliva tests were excluded from the features, resulting in R^2^ = 0.6963, only a small decrease compared to R^2^ = 0.7010 when all items were used, which was not much difference (Figure 3). The im-portance of the features (blood test items) was then calculated using the Feature_Importance_ parameter. The least important items were progressively excluded and continued to be evaluated accordingly. R^2^ remained the same in performance with R^2^ = 0.6937 when the number of feature items decreased to 15, slightly decreasing to R^2^ = 0.6570 when the number of features decreased to 10 and to R^2^ = 0.5673 when the number of features decreased to 5 The number of features decreased to 5. The top 15 features judged to be the most important are listed in Table 2.

**Table 2.** Top fifteen feature items ranked by importance.

| Feature | Importance |
| --- | --- |
| HBA1c | 0.163994 |
| ferritin | 0.134489 |
| free cholesterol | 0.115059 |
| urea nitrogen (BUN) | 0.095047 |
| albumin (ALB) | 0.075884 |
| alkaline phosphatase (ALP) | 0.071181 |
| mean corpuscular volume (MCV) | 0.046920 |
| red blood cell count (RBC) | 0.045560 |
| copper (Cu) | 0.043293 |
| blood sugar (BS) | 0.041645 |
| free fatty acids (FFA) | 0.036925 |
| alpha 2 globulin (A2G) | 0.035531 |
| platelet count (PLT) | 0.035518 |
| triglycerides (TG) | 0.029847 |
| lactate dehydrogenase (LDH) | 0.029100 |

**Figure 3.**
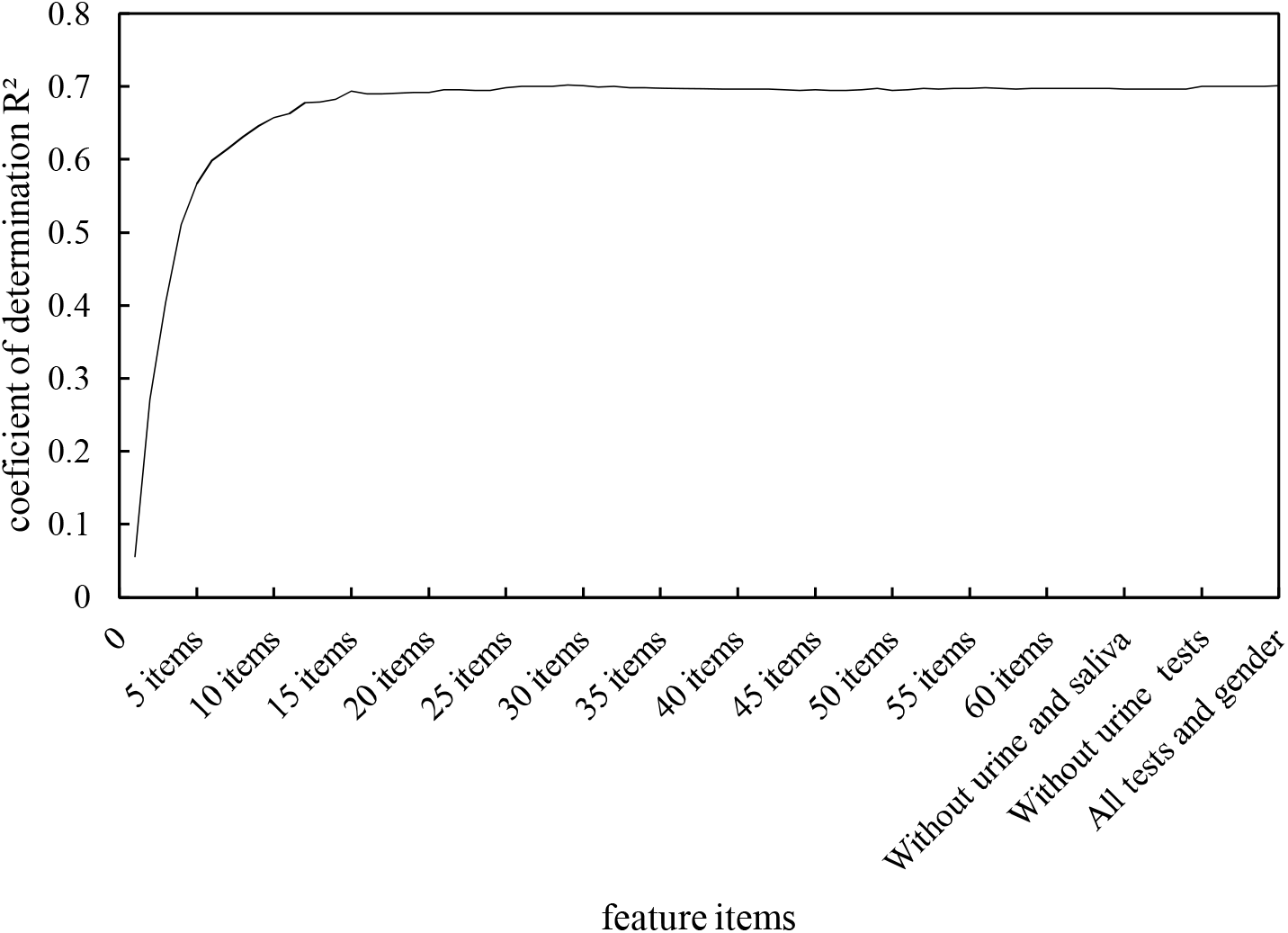
Coefficient of determination (R^2^) versus the number of feature items for model training. R^2^ slightly decreases from 0.7010 using all features to 0.6963 with only blood tests, and declines further as the least important blood test features are removed, down to 0.5673 with 5 features.

### 3.3. gender differences

After splitting the data into 80% (9,243) as training data sets and 20% (2,311) as test data sets, the data were evaluated separately for men and women. Results revealed similar performance for both genders: R^2^ = 0.711 for men and R^2^ = 0.695 for women.

In our study, we aimed to determine if there exists a discernible age estimation difference between postmenopausal and menstruating women, which was prompted by a subtle distinction observed in the scatterplot analysis for women around 50s (Figure 4). To verify this, we employed a T-test comparing the estimated ages of the two groups and found a significant age estimation disparity, with postmenopausal women being generally rated as older (P-value = 4.970E-50). Moreover, the median estimated age comparison between 45 and 55 years showed that postmenopausal women were, on average, estimated 5.22 years older (Table 3).

**Table 3.** Median estimated age per actual age for menstruating and menopausal women.

| Actual age (y.o.) |  | 45 | 46 | 47 | 48 | 49 | 50 | 51 | 52 | 53 | 54 | 55 |
| --- | --- | --- | --- | --- | --- | --- | --- | --- | --- | --- | --- | --- |
| Median estimated age (y.o.) | Menstruating women | 41.76 | 42.87 | 44.73 | 44.09 | 45.00 | 45.76 | 46.08 | 48.71 | 50.87 | 55.06 | 56.95 |
|  | Menopausal women | 47.45 | 54.94 | 42.38 | 53.60 | 54.32 | 50.09 | 52.55 | 54.85 | 54.82 | 57.72 | 56.59 |

**Figure 4.**
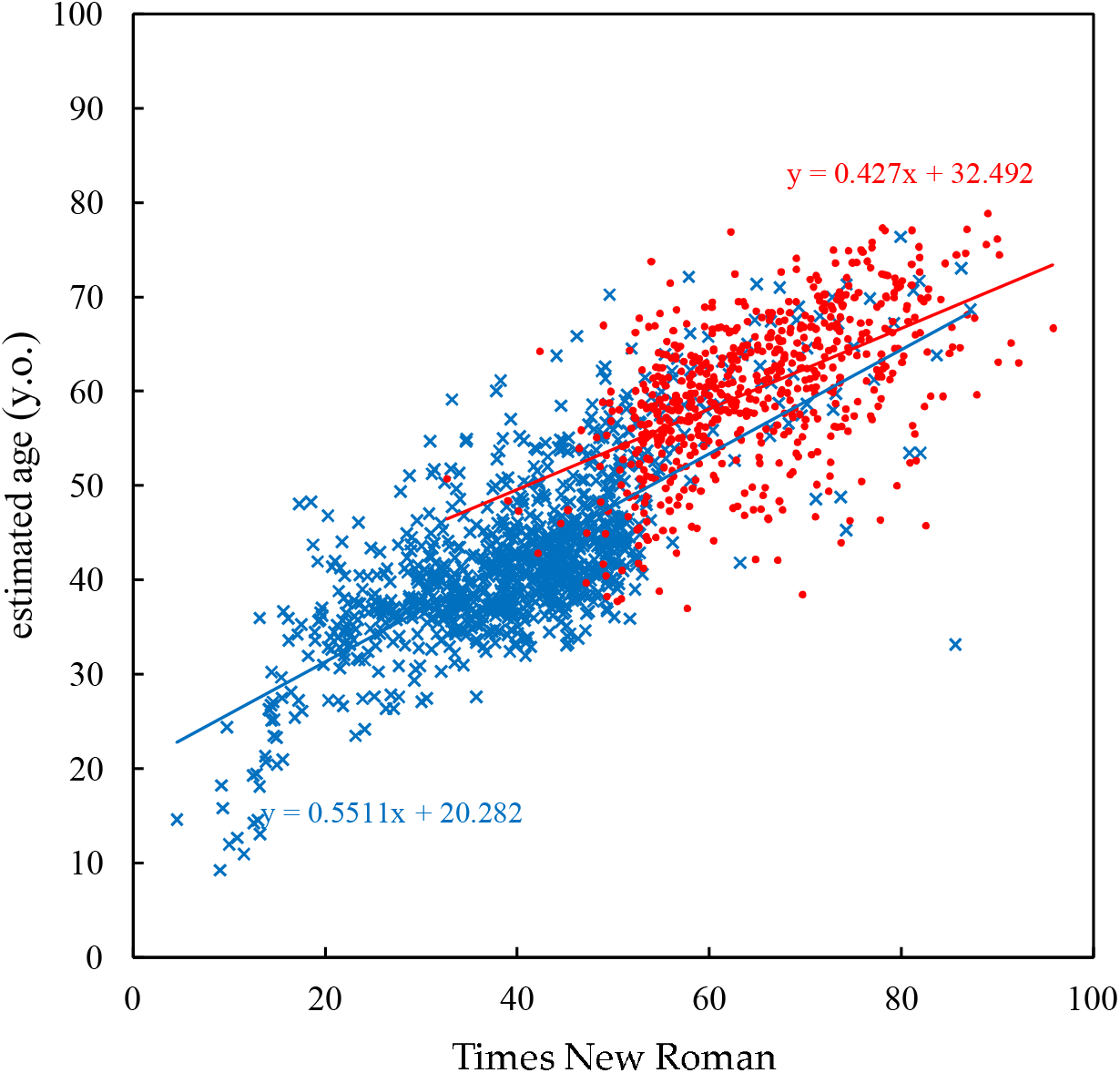
Scatterplot of actual age versus estimated age using a random forest model for menstruating women (blue) and postmenopausal women (red). The plot reveals a significant age estimation difference around the 50s age range, with the regression lines demonstrating that menstruating women are generally estimated to be younger than postmenopausal women.

## 4. Discussion

The results of this study suggest the possibility of using blood test results to estimate age. Specifically, the completeness of the training data sets significantly influenced the estimation’s accuracy. The selection of features in blood tests was also important, particularly HBA1c, ferritin, free cholesterol, urea nitrogen (BUN) and alkaline phosphatase (ALP), which were shown to be useful for age estimation. As for HBA1c, it is an important indicator of diabetic complications and is reported to be strongly related to ageing, even in non-diabetic individuals [21]. Ferritin has also been reported to be associated with ageing and its link to brain dysfunction in older people [22-24]. As for gender, it was considered to be an important feature in age estimates, as indicated by the data on gender differences in life expectancy. However, gender ranked 51st in importance overall. This does not necessarily imply that gender is less important, but rather that the overlap of gender information in blood data reduces the need to include gender itself as an explanatory variable. In fact, gender can be estimated well from blood data using the random forest method by same 15 item datasets as in 3.2. Evaluation of age estimation (accuracy = 0.906534).

In the future, it is conceivable that highly accurate age estimation could be achieved by adding other data sets, such as physical information, which were not used in the pre-sent analysis. However, various factors, including lifestyle choices like personal habits, diet, exercise, stress, medical history, genetics, and overall health, might affect the correlation between blood age and actual age. Therefore, direct application of these estimation results to health assessment and disease risk prediction requires deeper understanding and research. This is essential to ensure that the influence of these external factors and variables is accurately understood and incorporated into models. Furthermore, a detailed investigation into the causes of individual cases where discrepancies between the estimated age from blood test results and actual age were observed would provide a clearer understanding of the relationship between estimated age and health status. Gender differences were also observed, with the variation in estimated age among post-menopausal women in particular being an interesting phenomenon, it is possible that hormonal fluctuations and metabolic differences may be influencing this [16]. Future research should aim to elucidate the biological mechanisms behind these differences in detail.

## 5. Conclusions

This study attempted to estimate the age of individuals through blood test results and suggested that this could bring new perspectives to the field of preventive medicine. Improved estimation accuracy and the use of large data sets could contribute to individual health status assessment and disease risk prediction. As we look ahead, it’s vital to explore the complex interactions between blood age, lifestyle, genetics, disease, and other variables, to harness the full potential of this approach in preventive medicine.

## Author Contributions

The study was initiated by SK and MK to assess the validity of blood test data for human health problems; YS supported them as an expert in nutrition and blood parameters; OY provided advice on AI-based statistical analysis as an expert; SI and KH were primarily involved in the analysis and interpretation of the results from a medical point of view. The manuscript was first drafted by SK, including figures and tables, to which the other co-authors made additions and corrections. All co-authors participated in the final completion of the article.

## Funding

This research received no external funding.

## Institutional Review Board Statement

The study was conducted according to the guidelines of the Declaration of Helsinki and approved by the Research Ethics Committee of Orthomolecular Nutrition Laboratory (reference number: 2024_1-1; approved date: 24 Jan. 2024).

## Informed Consent Statement

I Informed consent to use the data for research purposes was obtained from the participants at the time of screening tests.

## Data Availability Statement

The data sets analyzed during the current study are available from the corresponding author on reasonable request.

## Conflicts of Interest

The data used in this study were received from KYB Medical Service Co., Ltd. SK, MK, and YS are affiliated with KYB Medical Service Co., Ltd.

## Notes

### Funding Statement

This study did not receive any funding.

### Author Declarations

Approved by Orthomolecular Nutrition Laboratory Research Ethics Review Committee.

